# Design and user experience testing of a polygenic score report: a qualitative study of prospective users

**DOI:** 10.1101/2021.04.14.21255397

**Authors:** Deanna G. Brockman, Lia Petronio, Jacqueline S. Dron, Bum Chul Kwon, Trish Vosburg, Lisa Nip, Andrew Tang, Mary O’Reilly, Niall Lennon, Bang Wong, Kenney Ng, Katherine H. Huang, Akl C. Fahed, Amit V. Khera

## Abstract

**Background:** Polygenic scores – which quantify inherited risk by integrating information from many common sites of DNA variation – may enable a tailored approach to clinical medicine. However, alongside considerable enthusiasm, we and others have highlighted a lack of systematic approaches for score disclosure. Here, we review the landscape of polygenic score reporting and describe a generalizable approach for development of polygenic score disclosure tools for coronary artery disease.

**Methods:** First, we assembled a working group of clinicians, geneticists, data visualization specialists, and software developers. The group reviewed existing polygenic score reports and then designed a two-page mock polygenic score report for coronary artery disease. We then conducted a qualitative user-experience study with this report and an interview guide focused on comprehension, experience, and attitudes. Interviews were transcribed and thematically analyzed for themes identification.

**Results:** We conducted interviews with ten adult individuals (50% females, 70% without prior genetic testing experience, age range 20 to 70 years) recruited via an online platform. We identified three themes from interviews: (1) visual elements, such as color and simple graphics, enable participants to interpret, relate to, and contextualize their polygenic score, (2) word-based descriptions of risk and polygenic scores presented as percentiles were most often recognized and understood, (3) participants had varying levels of interest in understanding complex genomic information and therefore would benefit from additional resources that can adapt to their individual needs in real time. In response to user feedback, colors used for communicating risk were modified to minimize unintended color associations and odds ratios were removed. Of note, all 10 participants expressed interest in receiving this report based on their personal genomic information.

**Conclusions:** Our findings describe a generalizable approach to develop and test a polygenic score disclosure tool that is desired by the general public. These results are likely to inform ongoing efforts related to polygenic score disclosure within clinical practice.

## Background

In a new era focused on preventive genomic medicine, polygenic scores have gained traction in clinical medicine as a method for stratifying individuals by disease risk – an approach that can enable targeted interventions [1–3]. This has led several commercial and academic groups to launch efforts to compute and report polygenic scores for patients and consumers. However, approaches for developing and reporting polygenic scores have been highly variable [4,5]. While there have been efforts to standardize reporting of polygenic score development in scientific literature [6], best practice guidelines for clinical reporting of polygenic scores do not yet exist.

Most of our understanding on optimal methods to report genetic risk is based on monogenic diseases, which are only partially transportable to polygenic score reporting [7–12]. Polygenic score reporting is different from monogenic disease reporting in at least three ways: First, polygenic score results are calculated on the basis of hundreds to millions of single nucleotide polymorphisms and reported as continuous outcomes. Therefore, they cannot be summarized using Human Genome Variation Society (HGVS) nomenclature and classified by pathogenicity as is recommended for monogenic reports by the American College of Medical Genetics (ACMG), European Society of Human Genetics, and the Association for Clinical Genetic Science [13–16]. Further, Farmer et al., recommends the use of a neutral statement of fact to summarize genetic results on a report, such as ‘a change in gene XYZ was found’ [17]. However, since a polygenic score can only be calculated on the basis of genome-wide variation and interpreted in the context of comparison to a population, a parallel statement does not exist.

Second, recommendations for clinical management based on polygenic scores have not been fully characterized and are therefore not discussed in expert guidelines such as the National Comprehensive Cancer Network and the United States Preventive Services Task Force [18,19]. The absence of management guidelines based on polygenic scores is an obstacle for providing resources, references, and clinical guidance that both patients and clinicians value on a genomic test report [8]. Although the ACMG does not provide a specific recommendation to outline patient-specific recommendations and resources on a genomic result, they do state that it “is appropriate and helpful” [15].

Third, the potential utility of polygenic scores has most often been described in the context of risk stratification across a population and disease prevention rather than absolute risk prediction and diagnosis [20]. Given this shift in reporting scope and intended use, we need to better understand how numerical, text, and visual elements included in a polygenic score report influence the overall message of the report and patient perceived actionability of the information.

Recommendations for designing genetic test reports for both patients and non-genetics providers published by Farmer and colleagues provide a foundation for the creation of genetic test reports that expand beyond diagnosis of a monogenic disease [17]. However, the authors concluded that additional research is needed to tailor these reporting recommendations for polygenic scores.

We recognized an unmet need for research on design of polygenic score reports, risk disclosure tools, and accompanying educational resources that could be understood by both patients and non-expert clinicians. This gap in resources is particularly relevant given the increasing number of initiatives that aim to make polygenic scores accessible to many individuals at no to low costs through the United Kingdom Accelerating Detection of Disease Challenge, eMERGE network, direct-to-consumer genetic testing companies, and independent academic organizations [21,22]. The vast scope of these projects will require new tools for polygenic score disclosure and education through new service delivery models that may rely on novel media for communicating with patients and consumers.

Here, we describe a generalizable approach for design of polygenic score reports using a user-centric approach, conduct a qualitative research study of user understanding and experience, and discuss new media that could enable a personalized and interactive experience with results from polygenic risk models.

## Methods

### Working group for polygenic score report for coronary artery disease

We selected coronary artery disease (CAD) as a representative example of an important complex disease where polygenic scores have been well-validated with respect to risk stratification [23–26]. We assembled a working group that consisted of 2 cardiologists experienced with treating coronary artery disease and genetics research, 1 genetic counselor, 2 clinical genetic laboratory experts, 2 designers, and 1 software developer. Over a period of 6 months, members of the group worked on iterative design and content creation (Fig. 1).

**Fig. 1.**
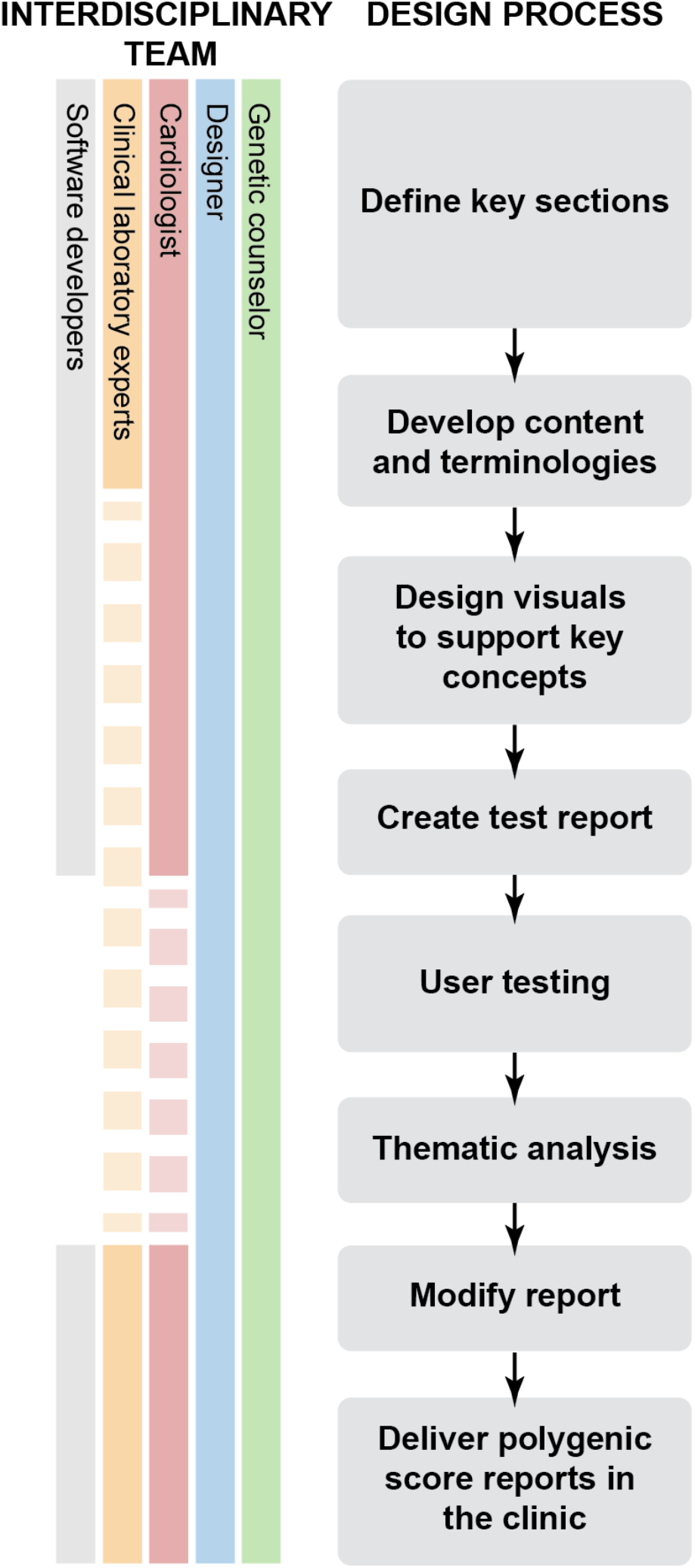
Developing a user-centered polygenic score report. An interdisciplinary team adopted a multi-step approach to create and iterate on a polygenic score report for coronary artery disease through a review of existing polygenic score reports and qualitative research methods.

### Review of the current landscape of polygenic score reports

We first reviewed publicly available polygenic score reports, identified through PubMed, internet search, the Polygenic Score Catalog [27], or provided through personal communication (Fig. 2). Reports were reviewed for (1) platform for risk disclosure (report medium), (2) personalized numeric risk estimate, (3) color schemes used, and (4) recommendations and resources provided.

**Fig. 2.**
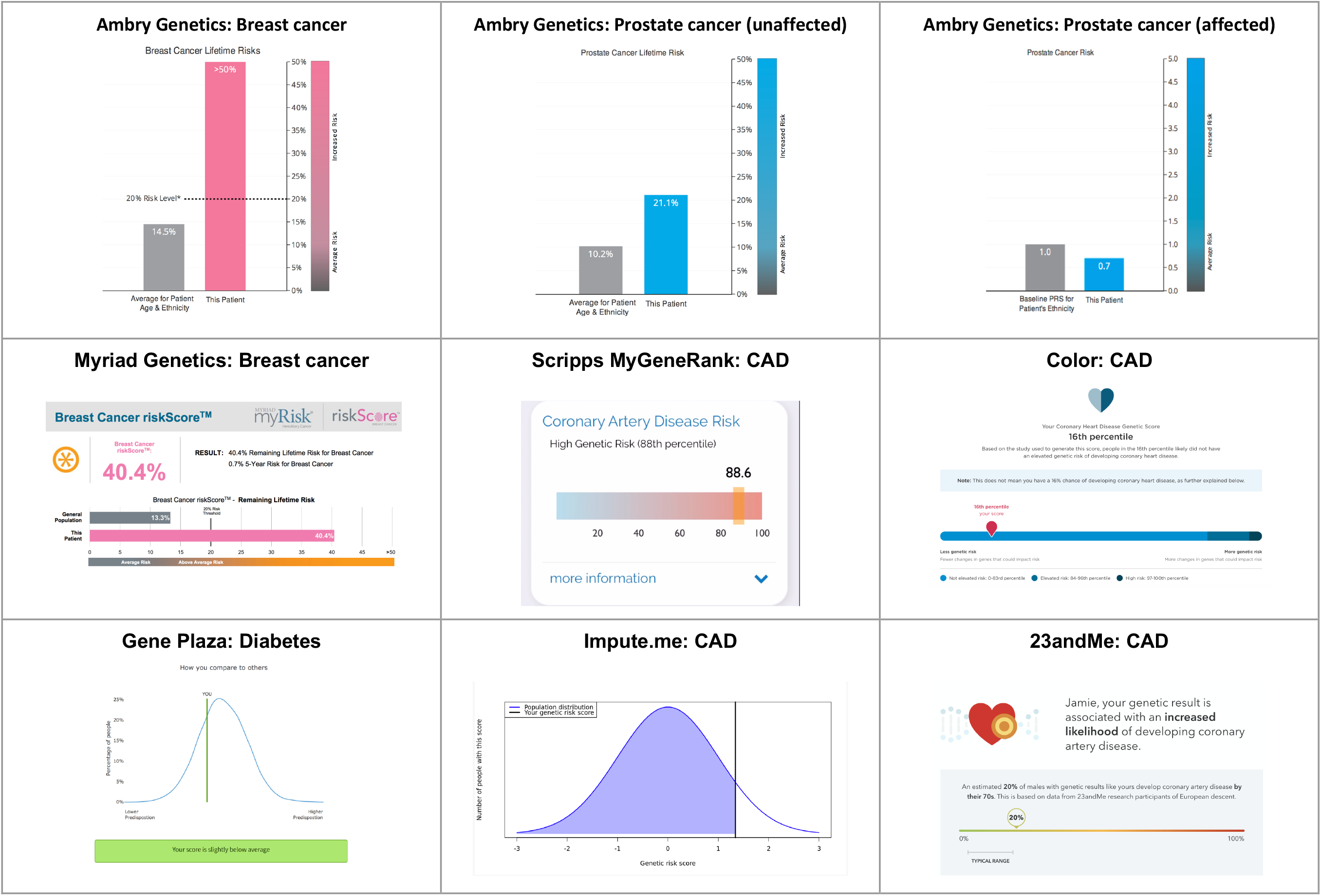
Polygenic risk score report visuals. Polygenic risk scores were compared on the basis of numeric estimates reported, risk description and colors used. ‘CAD’ represents coronary artery disease.

### Development of the polygenic score report

We developed a two-page polygenic score report that could be easily understood by both the general public and clinicians (Fig. 3). The report consisted of five sections, each intended to explain a major concept to support polygenic score interpretation and application for coronary artery disease, aided by a visual representation: (1) Information about the participant, (2) Participant’s score, (3) ‘What is a polygenic score?’ (4) ‘What is coronary artery disease?’ and (5) ‘How can I reduce my risk of coronary artery disease?’. Additionally, the report included a “Frequently Asked Questions” section to address common questions, provide a technical explanation of the test, and link to external resources.

**Fig. 3.**
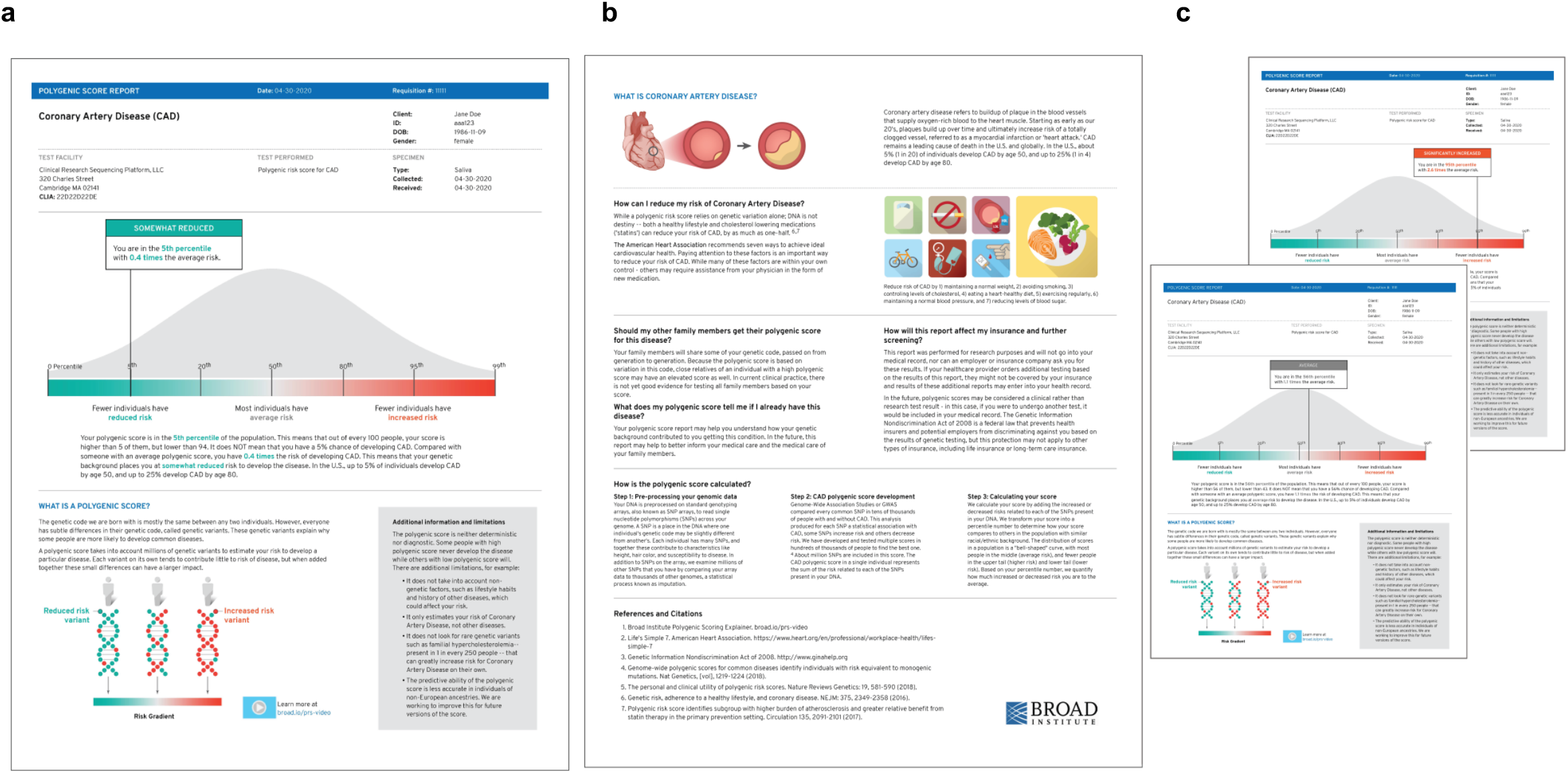
Mock polygenic score reports for coronary artery disease. **a**. Page one of 5th percentile (significantly reduced risk) mock report. **b**. Page two of all reports. **c**. Page one of 95th percentile (significantly increased risk) and 56th percentile (average risk) mock reports

### User experience of polygenic score risk disclosure tool

Ten participants were recruited through UserInterviews.com, an online recruitment and scheduling platform using quota sampling to enroll a relatively equal number of individuals by gender, age, and education. Individuals were required to be 18 years of age or older, have English as their native language, and be residents of the United States since disease prevalence described in the report was specific to the United States population. Eligible participants were excluded if they reported a personal history of coronary artery disease or worked in a genetics-related field.

We developed an interview guide consisting of 36 questions and prompts (Additional file 1) on the basis of preliminary feedback received from the working group’s colleagues, acquaintances, and family members. All participants provided consent to participate in the study, which involved both audio and video recording. Interviews were conducted by a visual designer trained in user experience research and a genetic counselor. Only the two facilitators and one participant were present for each session. Interviews were conducted virtually over Zoom video conferencing and lasted one hour in total. At the start of each session, participants were allowed up to fifteen minutes to independently review a mock polygenic score report for coronary artery disease indicating risk in either the 95th percentile (n= 4), 56th percentile (n=3) or 5th percentile (n=3). The facilitators then asked the participant to describe their experience viewing the report while being prompted to answer questions using the interview guide.

Each interview was recorded, manually transcribed by a member of the research team, coded and analyzed by the facilitators using thematic analysis [28]. Recurrent themes were identified from the data, coded, and summarized. This study was approved by the Mass General Brigham Institutional Review Board (2020P003088).

## Results

### Review of the reporting landscape for polygenic scores

We reviewed polygenic score reports from seven commercial and academic companies. Among these seven companies, coronary artery disease was the most commonly available polygenic score; others included type 2 diabetes and breast cancer. Reports were highly variable in terms of color, numeric risk estimate provided, categories used to describe amount of risk, and focus on resources and recommendations for the disease of interest (Fig. 2, Table 2)[2,29–41]. Three companies (Myriad, Ambry, 23andMe) provided at least one report that integrated a polygenic risk score with other clinical risk factors to generate an absolute percentage of risk.

Two companies (Ambry and Myriad) required a clinician to order the polygenic score. Of note, polygenic scores requiring clinician involvement were limited to individuals of ‘European ancestry’ (Table 2); an approach that is inconsistent with other clinical genetic tests. Both companies stated that the information provided on the report may be used to ‘better guide your patient’s medical management’ or ‘to assist in the development of a treatment plan’ [29,34]. All other reports were intended to be used only for research purposes and not to be used to guide medical management.

The remaining five companies allowed for a consumer-initiated assessment without the involvement of an individual’s healthcare provider. These consumer-initiated polygenic score assessments were calculated based on either genotype or sequencing data generated from a new saliva sample sent in by the consumer or raw genotype data shared directly with the company by the consumer. Results from consumer-initiated assessments were communicated via consumer-facing online portals or a phone application.

### Development of a new polygenic score report for coronary artery disease

Color was chosen as the main visual element to describe risk direction, using a scale from green, to grey, to red. This color scale was intended to leverage cultural associations of go-stop and cool-warm, with the grey serving as a neutral tone to represent average risk as the baseline for comparison. This color gradient was used across all risk-related visuals to bridge the relationship between an individual’s polygenic score with the genetic component from which it comes [42].

A risk score ‘flag’ containing the individual’s risk category, percentile, and odds ratio was positioned on the scale from significantly reduced risk (left) to significantly increased risk (right). The flag highlighted the risk category and was displayed prominently with a solid color-coded background of green, grey, or red. The body of the flag incorporated the same color-coding in the percentile and odds ratio text, calling attention to these values and reinforcing their association to risk direction. For this coronary artery disease polygenic score report, the working group chose to incorporate the following word-based descriptions to categorize risk: ‘significantly reduced risk’ (0-5th percentile), ‘reduced risk’ (6th-19th percentile), ‘average risk’ (20th-79th percentile), ‘increased risk’ (80th - 94th percentile), and ‘significantly increased risk’ (95th - 99th percentile). These categories were defined based on review of literature and expert consensus [43].

The population distribution graph behind the risk score ‘flag’ was included to support percentile understanding, emphasizing where most people fall on the polygenic score. Labels were also placed below the distribution to further describe how many people were in each risk category. The risk gradient shared the same x-axis as the population distribution and percentile markers for consistency and repetition.

We included a textual description of the individual’s polygenic score and an explanation of their percentile below the graph. The same color-coding was applied to words pertaining to percentile, odds ratio, and risk category for further repetition and consistency. We also included statistics on coronary disease prevalence to provide the reader with additional context about their risk.

One unique aspect of coronary artery disease is that regardless of an individual’s genetic background, a healthy lifestyle is associated with reduced risk of the disease [44]. To communicate positive lifestyle changes that individuals can undergo to mitigate an increased genetic risk, we used simple graphics to describe seven ideal cardiovascular health measures described by the American Heart Association as Life’s Simple 7: Managing blood pressure, controlling cholesterol, reducing blood sugar, exercising regularly, following a heart-healthy diet, maintaining normal weight, and stopping smoking [45]. The number of those metrics is a strong predictor of all-cause mortality and mortality from diseases of the circulatory system [43,44].

### User experience testing to optimizing a polygenic score risk disclosure tool for coronary artery disease

Interviews were conducted with ten participants (5 females, mean age 50.3 years, SD = 14.8, 7 without any prior genetic testing experience) (Table 1). All (10/10) participants expressed interest in receiving this report based on their own genomic data.

**Table 1.** Demographics of user experience testing participants (n=10).

| Characteristics | Participants (N = 10) |
| --- | --- |
| Average Age in years (range) | 50.3 (27-70) |
| Female, n (%) | 5 (50) |
| Self-reported race/ethnicity, n (%) |  |
| White | 4 (40) |
| Black | 3 (30) |
| Hispanic or Latino | 2 (20) |
| Asian | 1 (10) |
| Educational exposure, n (%) |  |
| Finished high school | 1 (10) |
| Some college | 4 (40) |
| Undergraduate degree | 2 (20) |
| Postgraduate degree | 3 (30) |
| Experience with genetic testing, n (%) |  |
| Yes | 3 (30) |
| No | 7 (70) |
| Job | Building Design & Construction, Receptionist, Attorney, Lecturer, Freelance Development, Retired, Organizer, Analyst, Exam Proctor, 'not reported' |
| Location | California, California, California, Florida, Georgia, Georgia, Massachusetts, Massachusetts, New York, New York, New York, Oregon |

**Table 2.**
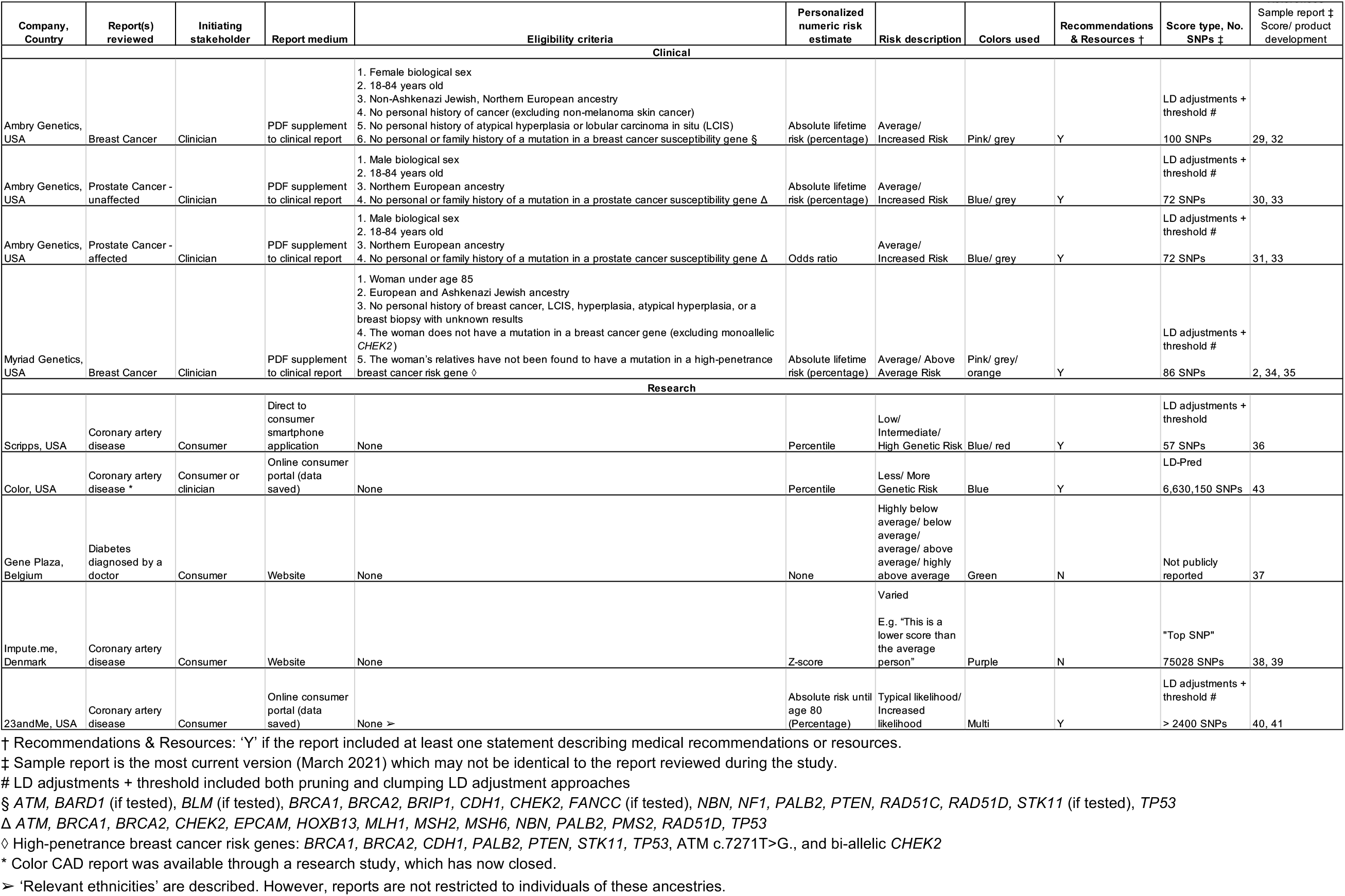
Landscape of polygenic score reports.

| Company, Country | Report(s) reviewed | Initiating stakeholder | Report medium | Eligibility criteria | Personalized numeric risk estimate | Risk description | Colors used | Recommendations & Resources † | Score type, No. SNPs ‡ | Sample report ‡ Score/ product development |
| --- | --- | --- | --- | --- | --- | --- | --- | --- | --- | --- |
| **Clinical** |  |  |  |  |  |  |  |  |  |  |
| Ambry Genetics, USA | Breast Cancer | Clinician | PDF supplement to clinical report | 1. Female biological sex<br>2. 18-84 years old<br>3. Non-Ashkenazi Jewish, Northern European ancestry<br>4. No personal history of cancer (excluding non-melanoma skin cancer)<br>5. No personal history of atypical hyperplasia or lobular carcinoma in situ (LCIS)<br>6. No personal or family history of a mutation in a breast cancer susceptibility gene § | Absolute lifetime risk (percentage) | Average/ Increased Risk | Pink/ grey | Y | LD adjustments + threshold #<br>100 SNPs | 29, 32 |
| Ambry Genetics, USA | Prostate Cancer - unaffected | Clinician | PDF supplement to clinical report | 1. Male biological sex<br>2. 18-84 years old<br>3. Northern European ancestry<br>4. No personal or family history of a mutation in a prostate cancer susceptibility gene Δ | Absolute lifetime risk (percentage) | Average/ Increased Risk | Blue/ grey | Y | LD adjustments + threshold #<br>72 SNPs | 30, 33 |
| Ambry Genetics, USA | Prostate Cancer - affected | Clinician | PDF supplement to clinical report | 1. Male biological sex<br>2. 18-84 years old<br>3. Northern European ancestry<br>4. No personal or family history of a mutation in a prostate cancer susceptibility gene Δ | Odds ratio | Average/ Increased Risk | Blue/ grey | Y | LD adjustments + threshold #<br>72 SNPs | 31, 33 |
| Myriad Genetics, USA | Breast Cancer | Clinician | PDF supplement to clinical report | 1. Woman under age 85<br>2. European and Ashkenazi Jewish ancestry<br>3. No personal history of breast cancer, LCIS, hyperplasia, atypical hyperplasia, or a breast biopsy with unknown results<br>4. The woman does not have a mutation in a breast cancer gene (excluding monoallelic *CHEK2*)<br>5. The woman's relatives have not been found to have a mutation in a high-penetrance breast cancer risk gene ◇ | Absolute lifetime risk (percentage) | Average/ Above Average Risk | Pink/ grey/ orange | Y | LD adjustments + threshold #<br>86 SNPs | 2, 34, 35 |
| **Research** |  |  |  |  |  |  |  |  |  |  |
| Scripps, USA | Coronary artery disease | Consumer | Direct to consumer smartphone application | None | Percentile | Low/ Intermediate/ High Genetic Risk | Blue/ red | Y | LD adjustments + threshold<br>57 SNPs | 36 |
| Color, USA | Coronary artery disease * | Consumer or clinician | Online consumer portal (data saved) | None | Percentile | Less/ More Genetic Risk | Blue | Y | LD-Pred<br>6,630,150 SNPs | 43 |
| Gene Plaza, Belgium | Diabetes diagnosed by a doctor | Consumer | Website | None | None | Highly below average/ below average/ average/ above average/ highly above average | Green | N | Not publicly reported | 37 |
| Impute.me, Denmark | Coronary artery disease | Consumer | Website | None | Z-score | Varied<br>E.g. "This is a lower score than the average person" | Purple | N | "Top SNP"<br>75028 SNPs | 38, 39 |
| 23andMe, USA | Coronary artery disease | Consumer | Online consumer portal (data saved) | None ➤ | Absolute risk until age 80 (Percentage) | Typical likelihood/ Increased likelihood | Multi | Y | LD adjustments + threshold #<br>> 2400 SNPs | 40, 41 |

† Recommendations & Resources: 'Y' if the report included at least one statement describing medical recommendations or resources.
‡ Sample report is the most current version (March 2021) which may not be identical to the report reviewed during the study.

We identified three themes from coded transcripts: (1) Visual elements, such as color and simple graphics, enable participants to interpret, relate to, and contextualize their polygenic score, (2) Word-based descriptions of risk and polygenic scores presented as percentiles were most often recognized and understood by participants, (3) Participants had varying levels of interest in understanding complex medical and genomic information and therefore would benefit from resources that can adapt to their individual needs in real time.

## Theme One: Visual elements, such as color and simple graphics, enable participants to interpret, relate to, and contextualize their polygenic score

### 1. Color

Color was the predominant design element that influenced participants’ level of concern about their hypothetical genetic risk. As intended, participants successfully used color to interpret their report, and frequently referenced these color ‘zones’ in terms of bad/good, safe/danger, unanimously considering them to be intuitive and discussing their score in terms of where on the color gradient they are (Fig. 4a).

**Fig. 4.**
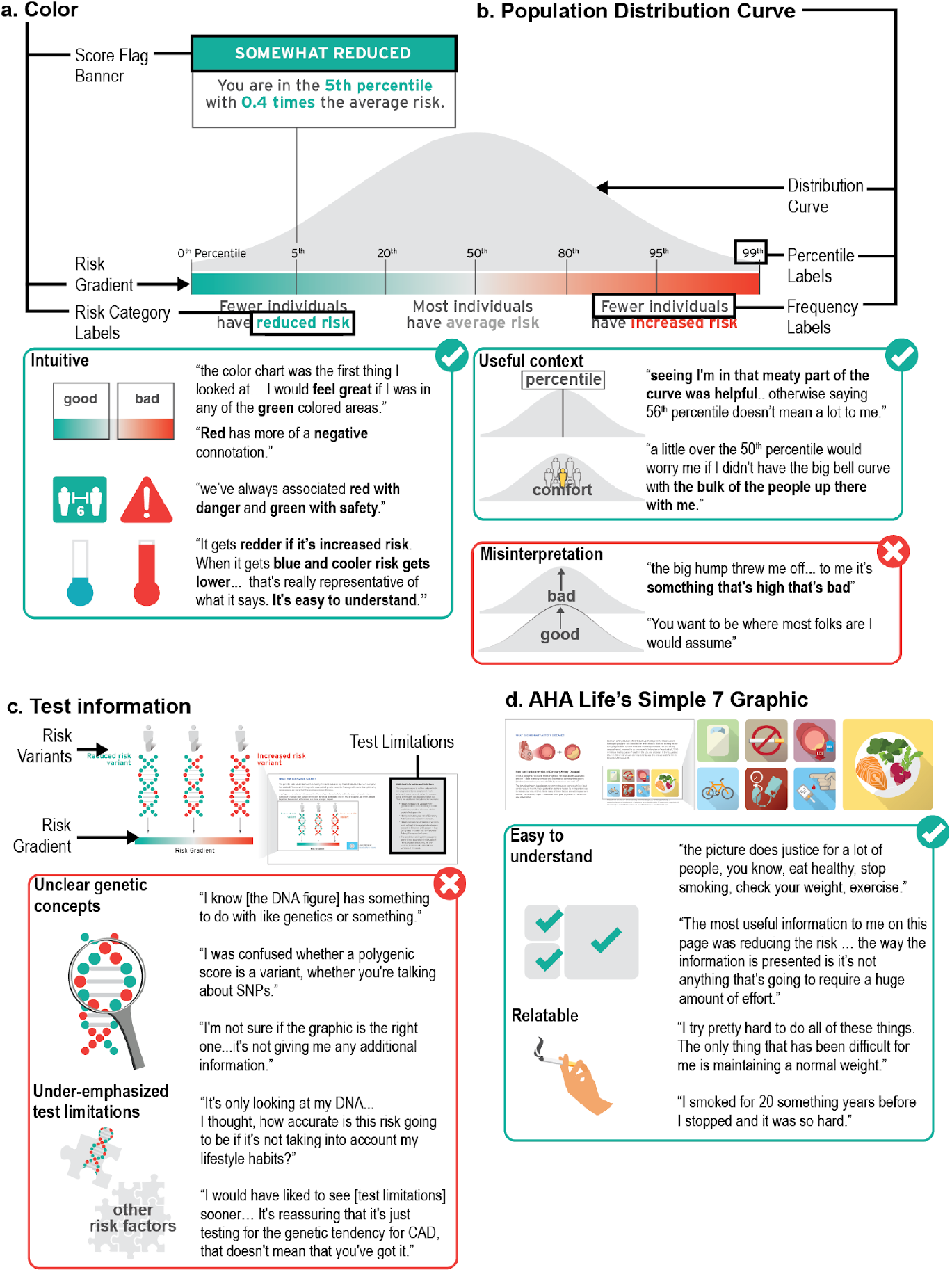
User experience testing results: Theme one. Visual elements, such as color and simple graphics, enable participants to interpret, relate to, and contextualize their polygenic score. **a**. Color was the predominant design element that influenced participants’ level of concern about their hypothetical genetic risk. **b**. Participants expressed differences in their understanding of the population distribution curve, interpretation of the underlying data, and association to its meaning. **c**. Participants were often unclear on genetic concepts and felt that test limitations were underemphasized. **d**. Participants found the cardiology and lifestyle graphics to be recognizable, relatable, and helpful for understanding the topic of the risk disclosure tool.

> *“I started with the chart, um, the color chart there, that was the first thing I looked at… So, looking at a score I would feel obviously great if I was in any of the green colored areas*.*”* (Participant 3)

> *“*… *we’ve always associated red with danger and green with safety*.*”* (Participant 10)
>
> *“When I see red, I think unsafe or something is not right*.*”* (Participant 2)

> *“*… *it gets redder, if it’s increased risk, I think that’s pretty good. I think it’s pretty expressive*… *when it gets blue [sic] and cooler, your risk gets lower down at the end, and it says, ‘reduced risk’, so I think that’s really good I think that’s really representative of what it says. It’s easy to understand*.*”* (Participant 6)

### 2. Population Distribution Curve

Participants expressed differences in their understanding of the population distribution curve, interpretation of the underlying data, and association to its meaning (Fig. 4b). One participant (9), who reported an undergraduate education level, was an outlier in their misinterpretation of the curve. This participant misinterpreted the height of the curve to represent the amount of risk, associating up with more risk/bad and down with less risk/good.

> *“the big hump kind of threw me off a little bit*… *to me it’s something that’s high that’s bad*.*”* (Participant 9)

However, other participants reported the curve on the risk disclosure tool to be helpful in their ability to interpret their score.

> *“It looks like most people are in the average section, like in the middle, like that’s why the curve is bigger there, cuz there are more people between the 20th and 80th percentile and then there’s less people with reduced risk and less people with increased risk based on their genetics*.*”* (Participant 1)
>
> *“the easiest to understand part of it is just the graphic itself… seeing I’m in that meaty part of the curve which generally, you know, says average and so I think that was helpful. … otherwise saying just someone’s in, you know, 56th percentile doesn’t really mean a lot to me*.*”* (Participant 3)

Some participants even found comfort in knowing how many other people were in the same group as them.

> *“You want to be where most of the folks are I would assume*.*”* (Participant 7)
>
> *“*..*even just being a little over the 50th percentile would worry me more if I didn’t have the big bell curve up there with the bulk of the people being up there with me*.*” “*…*I really like being in the middle*.*”* (Participant 6)
>
> *“most individuals have average risk, so if I’m going to be in the minority that has reduced risk I’m good with that*.*”* (Participant 4)

### 3. DNA Figure to Convey Test Information and Limitations

Participants’ recognition of the DNA figure often did not provide additional context for understanding polygenic risk variants (Fig 4c). For example, Participant 2 stated, “*I know [the DNA figure] has something to do with like genetics, genes or something*.*”* Another participant stated, “*I’m not sure if the graphic is the right one. I don’t think it’s - it’s not helping me; it’s not giving me any additional information or anything like that it’s just a different variance of the first graph*.” (Participant 7)

Other participants who were already familiar with genetic concepts found the graphic and supporting text to be useful, but still could not fully understand the concept of polygenic inheritance. For example, one participant said, *“I love explanations of how things work*…*it doesn’t mention SNPs here anywhere and I wonder if these were actually talking about SNPs, so I was confused whether a polygenic score is actually just a variant, whether you’re talking about SNPs*.*”* (Participant 6)

Regardless of whether participants understood the concepts being portrayed, they used color association to determine which individual they would be in the graphic based on their score. *“I really did not understand the picture if I’m being super honest. I understand as far as the color coordination from the top to the bottom the green, the gray, and the red. But I really didn’t understand…I don’t look at this and think DNA*.*”* (Participant 9)

However, it was the accompanying text about test limitations, shaded in grey, that enabled most participants to grasp the authors’ intended message for this section: this test only considers polygenic contributors to disease.

> *“I thought [the grey ‘limitations’ box] was very informative actually and it gives you a heads up that this [test] is not 100% guaranteed. It’s not taking into account if you’re a heavy drinker or heavy smoker or exerciser*.*”* (Participant 2)
>
> *“The test is only going to test you for coronary artery disease so any past problems that you have or that you have at the moment is not going to affect the test I guess*.*”* (Participant 5)

Importantly, one participant did not understand that this risk estimate did not consider lifestyle and past medical history.

> *“The information that I provided about my lifestyle and my personal statistics put me in the average range I would think that to get this [risk] information you would have to ask me a list of questions as far as my exercise activities, my past previous health scares, or surgeries*.*”* (Participant 9)

When participants were asked about their perception of the importance of information about test limitations, one participant responded, *“I would have liked to see [test limitations] sooner… It’s reassuring that it’s just testing for the genetic tendency for CAD, that doesn’t mean that you’ve got it*.*”* (Participant 6) Another participant noted that this section “cast a little doubt” about the utility of the test, stating that *“It’s only looking at my DNA, it’s not considering [lifestyle habits]. I thought, how accurate is this risk going to be if it’s not taking into account my lifestyle habits?”* (Participant 3)

### 4. Cardiology and Lifestyle Graphics

In general, participants found the cardiology and lifestyle graphics to be recognizable, relatable, and helpful for understanding the topic of the risk disclosure tool (Fig. 4d). When prompted to comment on their perception of the simple and familiar graphics of healthy lifestyle factors outlined by the American Heart Association, many participants took the opportunity to discuss their family medical history and their personal experience with these healthy lifestyle choices.

> *“I try pretty hard to do all of these things. The only thing that over my life has been difficult for me is maintaining a normal weight. But all the others I’ve been pretty good at*.*”* (Participant 10) *“… the picture does justice for a lot of people, you know, eat healthy, stop smoking, check your weight, exercise… My dad was 51 when he passed [from a heart attack] and as I get closer to that age I get more worried as well… I smoked for 20 something years before I stopped and it was so hard… you know, do I want to see my kids grow up?*… *And I did quit smoking*.*”* (Participant 5)

Additionally, participants felt that the lifestyle recommendations represented by these images felt manageable in terms of simplicity and cost.

> *“*… *since I’m [hypothetically] high risk I would definitely want to try to do anything I can to reduce that risk. And the ways that you guys provided is pretty simple, I would think, so like even by doing a few of those seven items it would hopefully help*.*”* (Participant 1)
>
> *“The most useful information to me on this page was reducing the risk … the way the information is presented is it’s not anything that’s going to require a huge amount of effort. … Eating better, doing some exercise, watching your cholesterol, all that stuff just seems doable and I think the way this information is presented here is like, oh maybe you do have this high-risk that you saw or high risk score, but, like, here are some great ways to help offset [the risk] I guess*.*”* (Participant 3)
>
> *“*… *it [lifestyle changes] seems easy, like you don’t have to spend thousands of dollars on, you can actually do it on your own. And it’s affordable*.*”* (Participant 2)

## Theme Two: Word-based descriptions of risk and polygenic scores presented as percentiles were most often recognized and understood by participants

Word-based descriptions of risk that provide high level explanations were frequently used by participants to summarize their takeaways from the risk disclosure tool (Fig. 5).

**Fig. 5.**
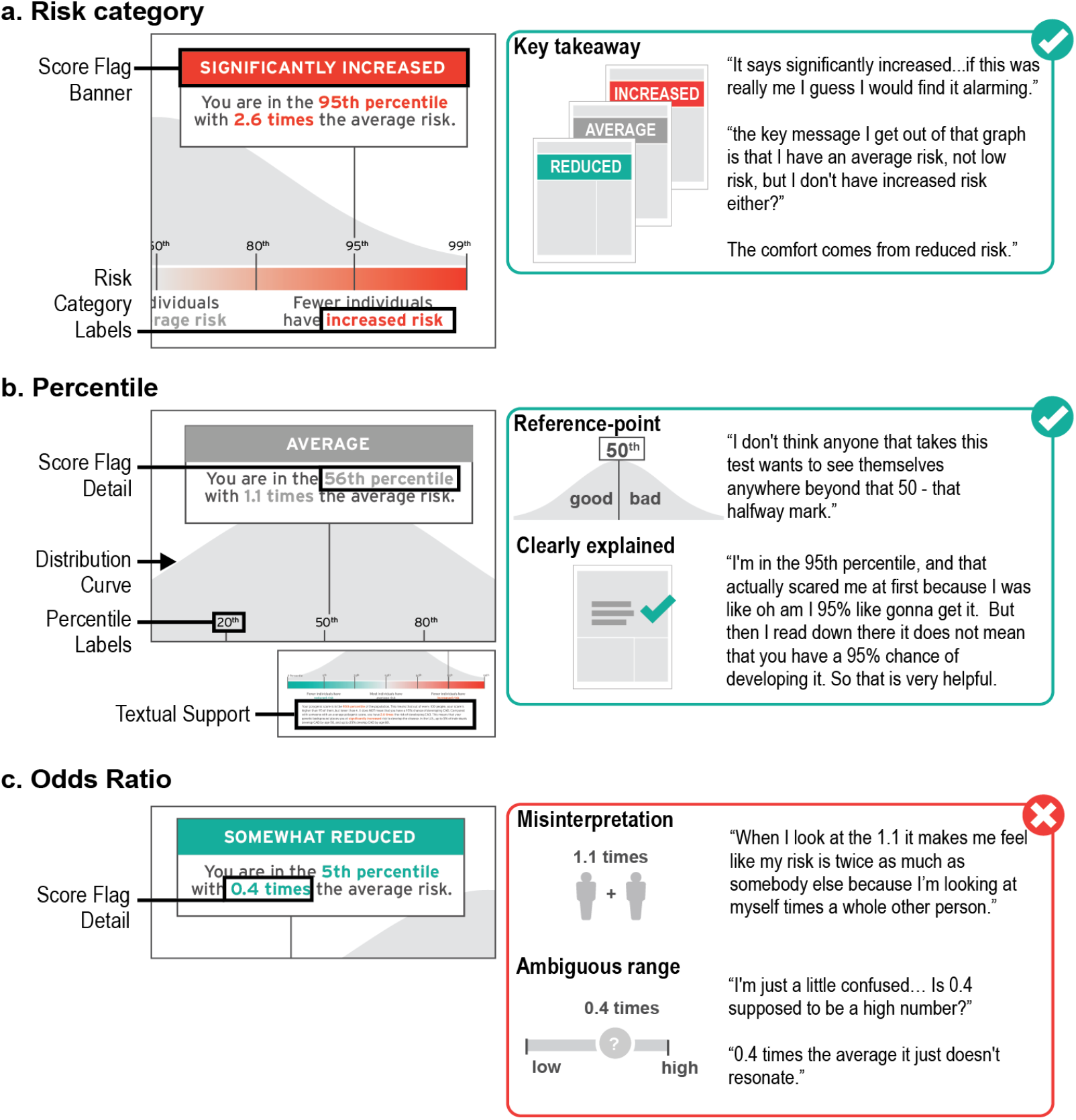
User experience testing results: Theme two. Word-based descriptions of risk and polygenic scores presented as a percentile were most often recognized and understood by participants. **a. ‘**Risk category’ is an interpretation of the numeric polygenic risk estimate. **b. ‘**Percentile’ is a polygenic risk estimate – on a scale from 0 to 100 – describing a participant’s location in a normal distribution. **c**. ‘Odds ratio’ is an estimate of risk that conveys magnitude of risk compared to ‘average risk’ of 1.0.

> *“So the key message I get out of that graph is that I have an average risk, I’m not low risk, but I don’t have an increased risk either?”* (Participant 3)

Importantly, this risk description was often a cause for concern or comfort for some participants.

> *“It says significantly increased*…*if this was really me I guess I would find it alarming*.*”* (Participant 10)
>
> *“The comfort comes from the reduced risk for me*.*”* (Participant 4)

When asked about information they found challenging in the ‘My Score’ section, many participants did not understand the odds ratio.

> *“When I look at the 1*.*1 it makes me feel like my risk is twice as much as somebody else because I’m looking at myself times a whole other person*.*”* (Participant 9)
>
> *“0*.*4 times the average it just doesn’t resonate really, but you know, having a reduced risk, even if you’re measured in the 5th percentile, that’s okay*. (Participant 4)

Some participants’ confusion surrounding odds ratio was rooted in not having a reference point for a ‘high’ and ‘low’ odds ratio.

> *“I’m just a little confused… Is 0*.*4 supposed to be a high number?”* (Participant 5)
>
> *“I was wondering why 2*.*6 times the risk instead of another number, like it’s really exact and I was just wondering how that came about*.*”* (Participant 1)

In contrast to an odds ratio with an ambiguous range of possible results, participants often discussed their familiarity with percentiles and commonly used percentile as the key reference point to interpret their score.

> *“I don’t think anyone that takes this test wants to see themselves anywhere beyond that 50 - that halfway mark*…*”* (Participant 7)
>
> *“*… *it says here that out of a hundred people your score is higher than 5 but lower than 94 which I mean is simple math*.*”* (Participant 5)
>
> *“I’m in the 95th percentile, and that actually scared me at first because I was like oh am I 95% like gonna get it. But then I read down there it does not mean that you have a 95% chance of developing it. So that is very helpful*.*”* (Participant 1)

Additionally, one participant acknowledged that neither number was most effective at communicating theoretical risk; rather, visual representations provided meaningful context for understanding all reported numerical values.

> *“You’re [in] the 95 percentile sounds significant… 2*.*6 times the average risk does not seem to be… the actual graph drives it home more so than the actual stats*.*” (Participant 7)*

## Theme Three: Participants had varying levels of interest in understanding complex medical and genomic information and therefore would benefit from resources that can adapt to their individual needs in real time

Universally, participants expressed their preference for receiving medical reports in simple terms with clear interpretation of results.

> *“*…*[medical reports] can become overwhelming and you know for me, I want you to get to the point, where do I stand, and how did you come up with this information, and how do I relate to everybody else in my class, and some of the things that I can possibly do*.*”* (Participant 10)

Nevertheless, participants had different preferences for the depth of information that they desired in this tool (Fig. 6); Participant 6 articulated that “*it would be great if you got a choice between”* the amount of information you receive. For example, some participants appreciated seeing a simple statistic for coronary artery disease prevalence.

**Fig. 6.**
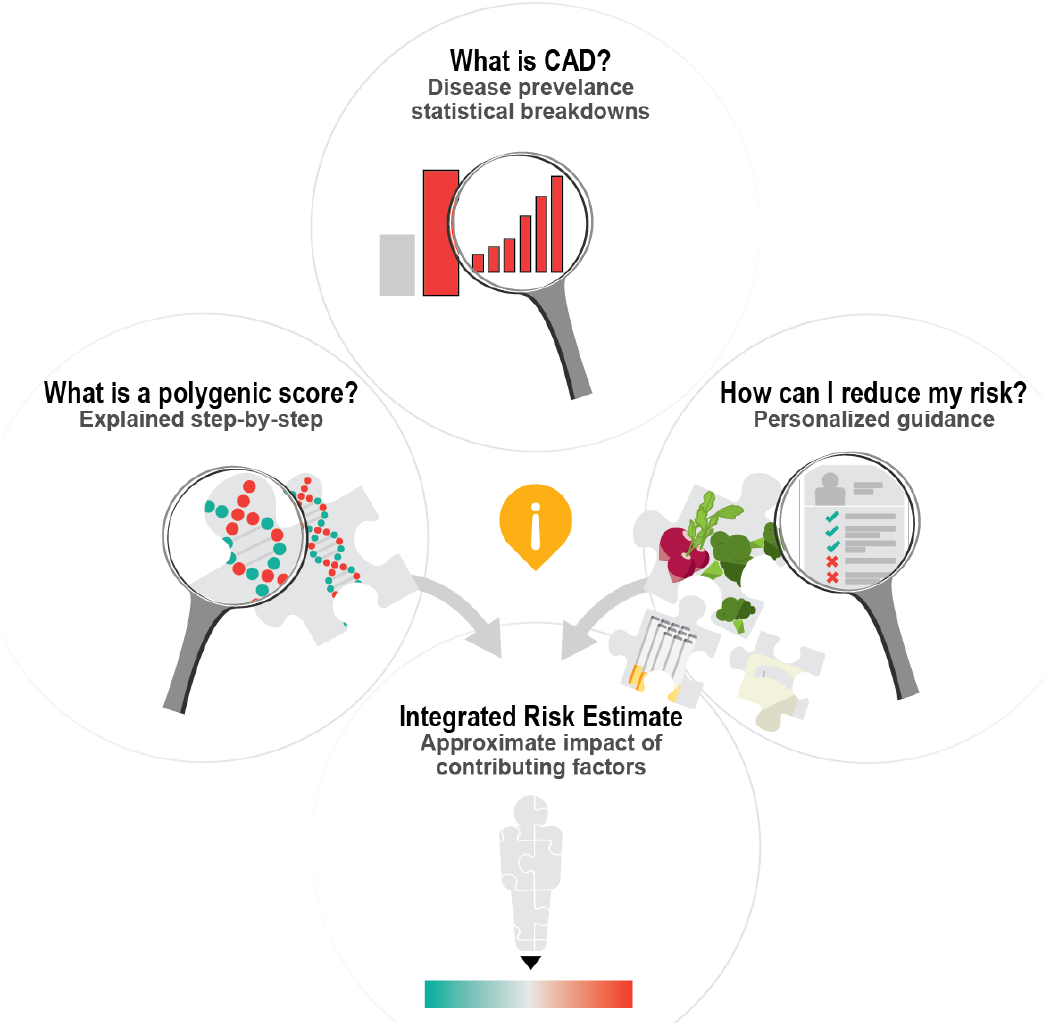
User experience testing results: Theme three. Participants had varying levels of interest in understanding complex medical and genomic information and therefore would benefit from resources that can adapt to their individual needs in real time.

> *“*…*1*.*1 times the average risk*…*that’s not that alarming, but when you say*… *people in your [age] group with this risk are up to 40% [at risk to] develop [CAD], that makes it a little bit more serious?” (Participant 3)*

While information about disease prevalence was included twice in the risk disclosure tool in sentence format, some participants shared that they were interested in receiving more detailed information about prevalence to further personalize the information.

> *“It said 1 in 20 [people will develop coronary artery disease] when you’re young, and then goes up to 1 and 4 when you’re like 80. But [I would like to see] more statistics about when you’re 65 or 70 broken down more*.*”* (Participant 6)

Other participants expressed confusion about the stated prevalence estimate, suggesting that additional explanation of disease prevalence, perhaps in a different format, is critical to minimize confusion and reduce the number of calculations on the individual’s part.

> ***“***… *depending on what your age is now, [your risk to develop coronary artery disease] could be from 1 in 5 [to] 1 in 25. So where do I fit in there?* … *I just dismiss that part okay, but* … *some people might just look at it and be like oh my God, I have a 25% chance …”* (Participant 9)

Another topic for which participants had variable interest in receiving more information was the ‘how to reduce your risk’ section. While some participants were satisfied with the amount of information in this section, many participants, particularly those that received ‘significantly increased risk’ mock score report, communicated that they would like additional guidance on how to adopt healthy lifestyle factors.

> *“this report isn’t going to detail a diet plan of course for anyone or an exercise plan for anyone, but I think there’s room for more information concerning a healthy lifestyle if they fall into a higher risk category. … I think a lot of folks just don’t know where to start*.*”* (Participant 7)
>
> *“I would love some dietary [suggestions] on how to reduce your cholesterol… some of these are complicated issues for people to deal with and giving some ideas on how to actually do proactive things would be really good*.*”* (Participant 6)

Other participants relayed that their interest in engaging with the risk disclosure tool may depend on the result. For example, one participant stated, *“if I was low I probably wouldn’t have read the whole thing*.*”* (Participant 1) Another participant suggested that some information reminded them of ‘medication inserts’, *“not very many people actually read those, but some people do. … you can ignore it if you want to, or you can go seek it out*.*”* (Participant 3) This raises the question of how best to tailor an individual’s experience viewing a genomic risk disclosure tool based on their individual result.

Some participants also indicated that other media, such as a website or a video, would be helpful for learning more about their score.

> *“… a couple videos that explain … What is coronary artery disease? What is the score? How do we calculate it? … Even if it’s just kind of a summary of the words that are there, I think having someone tell it to you is a lot easier than trying to read it and comprehend it*.*”* (Participant 3)
>
> *“I wouldn’t mind seeing like a little video clip of you know maybe a grandma and a granddaughter going to get this test done* … *you know actually see that it’s real live family members taking this test, and if it seems like it’s good results for them, then they could probably be good results for me*…*”* (Participant 2)

Participants also expressed their interest in learning more about how genetics and lifestyle choices together influence overall risk for coronary artery disease.

> *“*…*for example, out of 100 people, 80 of them who maintained the normal weight, avoided smoking, ate healthy, exercised, reduced sugar, they [saw] results in a matter of 30 days or 2 months, something of that nature. [This fact] makes it seem easy, like you don’t have to spend thousands of dollars on [them], you can actually do it on your own. And it’s affordable*.*”* (Participant 2)

Others described their interest in seeing a personalized overall risk estimate based on genetics and lifestyle factors.

> *“*…*Here is your risk score, and here is this other graphic that shows you can bring your risk down by 10 points or whatever if you were to exercise, and another three points if you were to eat right*.*”* (Participant 3)
>
> *“I specifically would be able to see the effects that my actions are having, because right now I can read this and it says that if I do these things my risk will be lower, but like how much lower? Is it worth me exercising, changing my diet, quitting smoking if in all actuality it’s only going to reduce my risk like 1 percentile? Or reducing it to whatever*… *I’m just saying: is all that effort worth it?”* (Participant 3)

Although not all participants felt that incorporating concrete numbers was critical for an interactive tool that layers lifestyle choices on genetic background. Rather, a direction and a broad estimate of magnitude would be sufficient.

> *“If you were to say okay, ‘do these 5 things in your risk goes from 56th to 35th percentile’*… *I don’t know what that means really. But if you were to say, ‘you’re going to go from this average risk to this, like, reduced risk, but your reduced risk is still at the higher end of the reduced’ -- then, yeah, I can get that a lot more*.*”* (Participant 3)

It was rare for participants to be completely satisfied with the information included in the risk report. Even though one participant stated that he received all of the information he “*need[ed] to know about polygenic scores in general”*, he still wanted more information about how valid and trustworthy the test is, especially “*if other people or other agencies or organizations have validated this particular test*.*”* (Participant 10)

At the end of the user experience session, participants were asked a series of questions about how interested they would be in receiving a report such as this one and how comfortable they would feel viewing this information on their own. All participants were interested in receiving a polygenic score for coronary artery disease, particularly those with a family history. Additionally, all participants indicated that they would be okay reviewing their polygenic score for coronary artery disease on their own; although, five stated that they would want to discuss it with their doctor at a later time for clarification and personalization of the information.

> *“I wouldn’t mind receiving it on my own going through it and then maybe printing it out and then my doctor going through it with me because there’s gonna be some big words there my doctor knows, you know break it down and let’s work up with a plan and how I can avoid or lessen or stop if possible whatever it is that’s negative like CAD*.*”* (Participant 2)

As one participant highlighted, *“a lot of people don’t go [to the doctor] often. Like twice a year. Maybe even once a year*.*”* (Participant 6) For these individuals, it will be crucial for companies and researchers to consider this as they develop polygenic score risk disclosure tools that are made available directly to patients and consumers.

### Modifying the report based on user feedback

Although color was well-understood by participants, we modified colors used to convey risk throughout the report to reduce the strength of association. The red-orange color, which was used to convey high risk, was altered to add cooler undertones; the green color, which was used to convey reduced risk, was changed to a blue/teal. In addition, we removed the polygenic score odds ratio from the risk flag to emphasize the numeric estimate that resonated most with users -- polygenic score percentile. Last, we created a video explainer to provide additional education and resources for users to learn more about polygenic scores [46].

## Discussion

In this qualitative study involving user experience testing of a polygenic score report for coronary artery disease, we describe a generalizable and reproducible process for designing a polygenic score report for a complex disease. Our findings highlight two key insights in genomic implementation of risk disclosure using polygenic scores, which provide important guidance for next steps (Fig. 7).

**Fig. 7.**
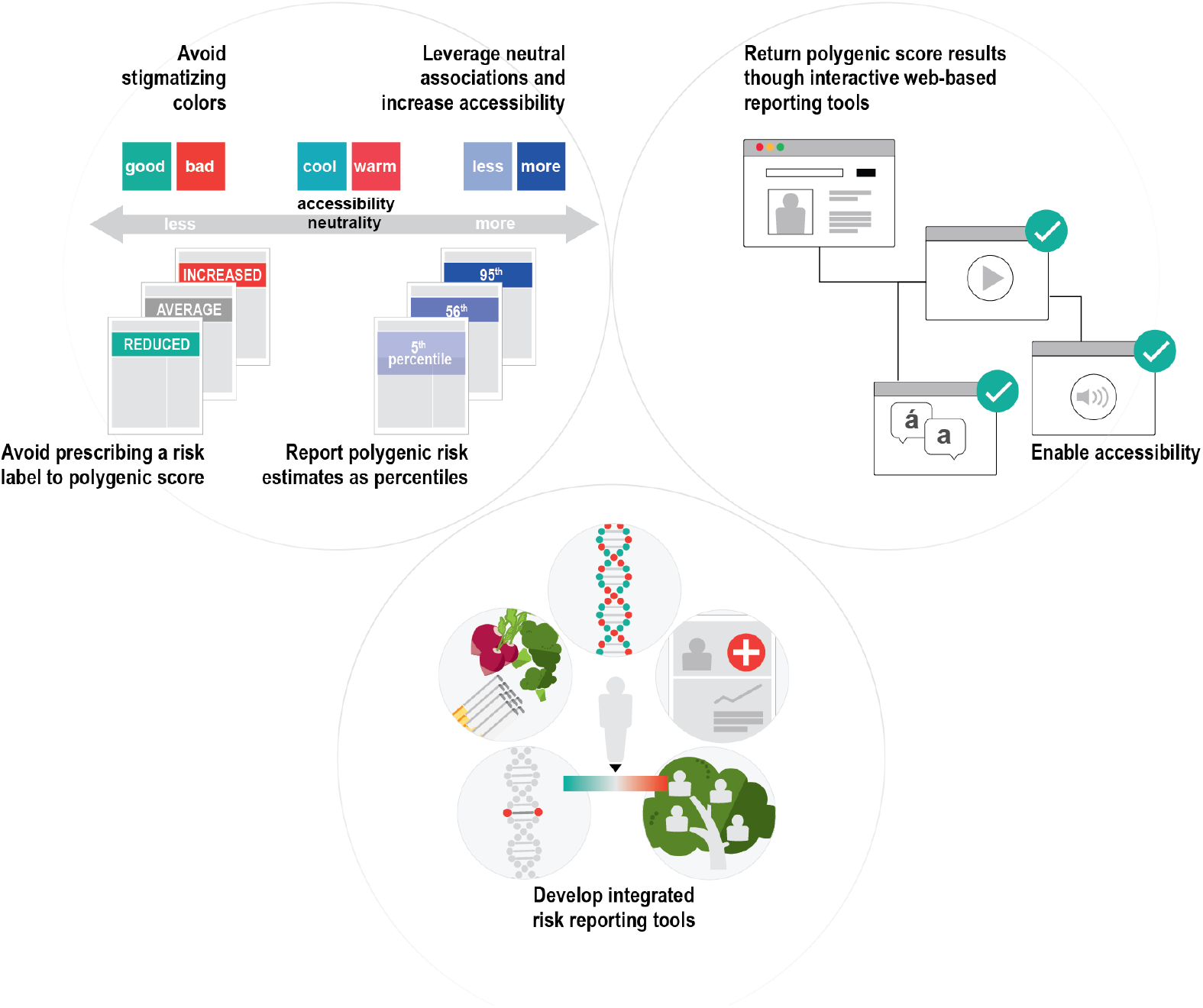
Next steps in genomic risk disclosure.

First, heterogeneity in the design of existing polygenic score reports and disclosure tools will present challenges for studying risk perception and clinical utility. A successful model for designing risk disclosure tools should take into consideration components of user-centric design and the intended utility of the risk model. Even seemingly routine design and reporting aspects of a report such as choice of color, visual and numerical estimates can have dramatic impact on understanding and perception of risk. For example, our study demonstrated that color, which varied across mock reports, was an important element of design that strongly influenced risk perception. The strength of participants’ red-green color association merits consideration of the intended goal for the use of color in the score report: enhancing readability, emphasizing key information, promoting perceived actionability, or sparking concern or comfort. For each of those goals, the choice of color would differ. In the case of the report in this study, the red-green color served strongly to spark concern or comfort.

Farmer et al. advises against the use of color to communicate results since the interpretation of color may vary from individual to individual. This recommendation is particularly relevant in our study given the lack of clarity surrounding clinical and personal utility of polygenic scores [47]. If color is used to convey the magnitude of disease risk and there are limited guidelines on how to mitigate this risk, we recommend that a clinician expert be made available to that patient or consumer to discuss this risk. Further, when selecting colors to convey information about health risks, we should be mindful of the inherent limitations of using color, such as the varying cultural associations of different colors, and its weakened impact for individuals with color blindness [48,49]. In future studies, it would be important to consider alternative options, such as a monochromatic color scale [50]. This could simplify the amount of visual-decoding required by patients, and would lend itself better to describing less/more risk, rather than low/high risk, a subtlety that nevertheless has implications for risk perception.

In addition to color, participants often recalled percentile and ‘risk category’ as the key messages of the report. Although percentiles do not correspond to medical guidelines at this time, we chose to emphasize a percentile as the main result since it provided an unambiguous range of reference -- 0 to 100 -- that was most understood by participants. Additionally, percentiles can be adjusted by genetic ancestry, self-reported race, and ethnicity. We removed the odds ratio from the report because it was poorly understood by participants and varies according to genetic ancestry, self-reported race, and ethnicity [51]. Before reporting a polygenic score as an odds ratio on a patient and clinician-facing report, the score should first be re-calibrated in a diverse cohort that is representative of the consumer or patient population receiving the score. Additionally, visuals that provide a simple explanation of an odds ratio should be included in this risk disclosure tool. An absolute risk estimate was not included in our report. At this time, we recognize that polygenic scores are less precise across populations and by integrating them with existing, problematic, race-corrected clinical risk models, we risk perpetuating and exacerbating health disparities [52,53].

To be transparent about the current limitations of polygenic scores for individuals of non-European ancestry, we included the following statement on page 1 of the report -- ‘The predictive ability of the polygenic score is less accurate in individuals of non-European ancestries. We are working to improve this for future versions of the score.’ One participant who self-identified as Black expressed concern about this limitation, but still recommended that we *“go ahead, roll [the polygenic score] out [to everyone] and just let the non-European ancestry [individuals] know it’s going to be a little long, plus or minus whatever variance*…*It’s probably still worth rolling it out to everyone*.” (Participant 7) A second participant who self-identified as Black also appreciated knowing this information and suggested that everyone be informed about this limitation before pursuing the test. Interestingly, this participant stated, *“put [the limitation about race, ethnicity, ancestry] on the reports that it pertains to… so if it does not pertain to your history that wouldn’t be any confusion. So if in the ethnicity part, when you’re doing the questionnaire, provide this information just to those that it pertains to*.” (Participant 9)

In the context of this study, we felt that it was appropriate to bin and assign risk categories by percentile to evaluate perceived value and meaning by participants. However, we appreciate the significance that these labels carry when clinical utility has yet to be established and when clinician experts are not involved in the return of these results. We encourage others in the field who are developing polygenic score reports and risk disclosure tools to recognize the significance of these labels for patients, consumers, non-expert clinicians, insurance companies, and proceed with caution.

Second, static genomic test reports that limit one’s ability to explore results are not ideal for polygenic risk disclosure. Risk disclosure tools that enable patients and consumers to have a personalized, interactive experience with their genomic report should be developed for polygenic score disclosure. One such medium to consider for this purpose is reader-driven narrative visualization, sometimes referred to as ‘Scrollytelling’. Reader-driven narrative visualizations are online, user-centric, interactive tools that enable dynamic storytelling through data visualization to educate the reader about complex concepts [54]. To date, this medium has been used to educate about public health issues such as the United States Measles outbreak in 2015 and the Covid-19 pandemic [55–57]. We see an unexplored opportunity to develop and evaluate the utility of reader-driven narrative visualization tools in the context of preventive genomic medicine to support personalized healthcare.

In the context of coronary artery disease, a reader-driven narrative visualization risk disclosure tool could provide patients with resources and guidance outside of physician-prescribed medications and tests. For example, some patients or consumers may benefit from additional resources or information about how to manage one’s cholesterol through diet. While a report may not be able to provide specific dietary recommendations, a report could consider referencing within the tool resources from the American Heart Association so that the patient can review them independently before discussing their result with a clinician. We anticipate that providing patients with the opportunity to be educated independently may also promote a more in-depth counseling experience between patients and their clinicians to discuss the individual’s motivation for pursuing particular healthy lifestyle changes and any barriers they are facing to adopt this change. Lastly, reader-driven narrative visualization tools for polygenic score results could also facilitate expansion of accessibility options for patients and consumers, such as providing quick language translation of content and enabling audio playback of content for individuals with visual impairments.

This study has several limitations. First, given the speed at which polygenic score reports are being developed and modified, reports may have been updated since our review of existing reports. Second, we aimed to recruit a cohort of participants with diverse backgrounds (education/work experience, age, geographic location, race/ethnicity) using a national online recruitment platform. However, our sample size was small and likely does not reflect the experiences of all individuals who may receive a polygenic score for coronary artery disease. Future studies should be conducted with more individuals in the general public with diverse backgrounds and individuals with a personal history of coronary artery disease.

Third, this study did not evaluate the clinicians’ experience viewing a polygenic score risk disclosure tool. It is important to acknowledge that clinicians may have different preferences for the design of a polygenic score risk disclosure tool than the general public, which could impede clear communication of information. As efforts for returning polygenic scores continue to advance, it will be important to assess perceived utility of report elements by clinicians with and without clinical genetics expertise, and to standardize terminology and thresholds for risk labeling by disease area.

Fourth, all participants stated that they would feel motivated to change their behavior if they received the mock report shown to them in this study. While intent is critical to the process of adopting healthy lifestyle behaviors, prospective studies are needed to assess behavior change based on receipt of actual polygenic score reports for coronary artery disease.

## Conclusions

Our qualitative study revealed that polygenic risk score disclosure presents a set of challenges that are not unique in genetics, but should be approached with additional prudence. This study demonstrated that polygenic score reporting tools, which promote informed and inclusive understanding of personal and family history was desired by participants.

Through personalized polygenic score reports that leverage dynamic, reader-driven narrative visualization approaches, there is a unique opportunity to pair genomic risk reporting with education, expand accessibility options, provide personalized health information and support, and ultimately empower individuals to stay healthy. Finally, there is a need for professional organizations to put forth guidelines and best practices for polygenic score disclosure and reporting. In particular, guidance is needed to standardize risk category nomenclature and corresponding percentile/odds ratio thresholds by disease area, numeric reporting requirements, information on score limitations, and disclaimers.

## Data Availability

For additional information about data referred to in the manuscript, contact D. Brockman.

## Acknowledgements

This study was funded by a sponsored research agreement from IBM Research, grant 1K08HG010155 (to A.V.K.) from the National Human Genome Research Institute, a Hassenfeld Scholar Award from Massachusetts General Hospital (to A.V.K.), a Merkin Institute Fellowship from the Broad Institute of MIT and Harvard (to A.V.K.).

## List of abbreviations

ACMG: American College of Medical Genetics

## Declarations

B.C.K. and K.N. are employees of IBM Research. A.C.F. is a consultant and holds equity in Goodpath. A.V.K. has served as a scientific advisor to Sanofi, Medicines Company, Maze Therapeutics, Navitor Pharmaceuticals, Sarepta Therapeutics, Verve Therapeutics, Amgen, Color, and Columbia University (NIH); received speaking fees from Illumina, MedGenome, Amgen, and the Novartis Institute for Biomedical Research; received sponsored research agreements from the Novartis Institute for Biomedical Research and IBM Research.

