## Additional file 1 for "Design and user experience testing of a polygenic score report: a qualitative study of prospective users"

**Supplementary Information. Interview Guide**

Thanks for taking the time to read through the report. For the user testing session we want to get a better sense about how you interpret and understand the information in the report so that we can make improvements to it. We'll go through each section of the report and I'll ask different questions to guide the discussion.

**General**

1. Can you walk me through the order of how you read in the report? / What was the first thing you looked at on the report?
2. After reading the whole report, can you tell me what the main thing this report is telling you?

**Risk Figure: Percentile**

3. What is the key message of this section?
4. What is your understanding about your risk to develop coronary artery disease?
5. What's your understanding of 'high risk/ average risk/ low risk'?
    a. What helped you to understand your risk?
6. What does your percentile tell you about your risk?
    a. What percentile do most people fall into?
    b. What percentile would you like to be in?
7. What information is the grey curve telling you?
    a. Does it help you understand your risk in any way?
8. What does the color coding tell you about your risk?
9. Did the labeling help you to understand the graphic?
10. Does this graphic leave you with any questions?

**Polygenic Score**

11. What was your main takeaway from this section?
12. Do you recognize this graphic?
13. What is this graphic telling you?
14. What is your understanding of a genetic variant?
15. What are the types of genetic variants?
16. What's the difference between an individual who is at *increased* risk and *reduced* risk in the graphic?
17. Did you read the text in the grey box? What is the information telling you? What was important? Was there anything that surprised you?
18. How does the information in the grey box help you to understand your score?

**Coronary artery disease/ Lifestyle**

19. Did you find the information in this section useful?
20. Using this report, can you tell how common is coronary artery disease?
    a. Does this feel common to you?
    b. Does this make you think differently about your risk , or make you any more or less concerned?
21. Do you feel like you could reduce your risk for coronary artery disease?
22. If you took this test again after improving your lifestyle, would you expect your results to be different?
23. How motivated would you feel to change your lifestyle?
24. How much of an impact do you think lifestyle could have on your risk?

25. Do any lifestyle factors seem harder or easier to do?
26. Do you think any lifestyle factors have a bigger impact on risk?
27. Would any additional information make you more likely to change your lifestyle?

**Report Structure**

28. How much effort do you feel is required to read and understand the document?
29. What was easy to understand? What information was difficult to understand?
30. How do you feel about the balance of text and visuals? Is there too much of one and not the other?
31. Was there any information or visuals that helped you understand the report?
32. Would a website or video be suitable complements or alternatives to this report?

**Other**

33. What is the maximum amount of money you would pay (max) for this report?
34. If you got this report, would you want to review this with your doctor?
35. This is a hypothetical report. How interested are you in receiving this report based on your actual genetic information?
36. Do you have anything else you would like to add/ share about your experience viewing this report?
